# Creating an Indexing Scheme for Case Series Articles

**DOI:** 10.64898/2025.12.19.25342712

**Authors:** Andrew Shahidehpour, Arthur W. Holt, Ang Michael Troy, Joe D. Menke, Neil R. Smalheiser

## Abstract

**Objectives:** Case reports and case series comprise a significant portion of the biomedical literature, yet unlike case reports, the National Library of Medicine does not index case series as a Publication Type. This hurts clinicians’ and researchers’ ability to retrieve, identify and analyze evidence from this type of study.

**Materials and Methods:** PubMed articles mentioning “case series” in title or abstract were characterized to learn what are considered to be case series by the authors themselves. We then set aside articles better indexed as other standard publication types – case reports, cohort studies, reviews and clinical trials -- as well as those that discuss (rather than report the results of) case series studies, to create a corpus of typical case series articles. A random sample of these articles was evaluated by two annotators who confirmed that the great majority satisfy a formal definition of “case series”.

**Results:** The corpus was utilized in an automated transformer-based machine learning indexing model. Case series performance of this model on hold-out data was excellent (precision = 0.887, recall = 0.952, F1 = 0.918, PR-AUC = 0.941) and manual evaluation of 100 articles tagged as “case series” revealed that 88% satisfied a formal definition of case series.

**Discussion and Conclusion:** This study demonstrates the feasibility of automatically indexing case series articles. Indexing should enhance their discoverability, and hence their medical value, for evidence synthesis groups as well as general users of the biomedical literature.

## Introduction

Case reports and case series are types of observational study designs which represent a significant part of the biomedical literature. Over 2.4 million case report articles are published and indexed within PubMed, and roughly 100,000 case series are published as well. Together, case reports and case series form the lowest tier of the Evidence Hierarchy in evidence-based medicine, but despite their lowly status, they contribute in important and unique ways to medical progress [1–3] and many systematic reviews include or specifically cover case reports and case series. Generally, case reports are unplanned eyewitness reports of one or a few patients, in contrast to case series which are generally planned analyses of a larger set of patients, generally four or more. Whereas Case Reports is a recognized Publication Type and indexed as such in MEDLINE and PubMed, no NLM indexing of case series articles exists at all, creating a gap in the ability of researchers to identify and analyze evidence from this type of study.

The traditional means of indexing articles involves deciding on a formal definition of “case series”, and training annotators to manually evaluate each article as it is published. Alternatively, one could create a manually annotated corpus of articles that can be used as a training set for automated machine learning methods [4–6], but a large amount of time and effort is required to train and manually annotate a large, representative gold standard corpus. As an alternative, we have pursued a data-driven semi-automated approach.

First, we aimed to characterize the overall body of PubMed articles that mention the phrase “case series” in the title or in the abstract. We hypothesize that articles which employ the phrase “case series” in the title should largely be articles that authors themselves feel are case series, or that discuss one or more case series (e.g., a review of case series on a given topic, or a discussion of proper design methodology). Articles mentioning “case series” only in the abstract may also include some case series studies, though we expect that they may often discuss case series or mention them incidentally. We acknowledge that authors are not always well-informed or correct about using the term “case series” [7–9], and in fact our analyses revealed many examples in which authors mislabeled their studies. Therefore, using these articles as a starting point, we aimed to remove the bulk of those articles which discuss (rather than report the results of) case series studies, as well as remove those better indexed as other standard publication types. This resulted in a corpus suitable to use as a training set for an machine learning model, which automatically indexes biomedical articles as Case Series when appropriate.

## Methods

### Overview

As shown in the flowchart (Figure 1), articles mentioning the phrase “case series” in title or abstract were retrieved from PubMed, processed, and progressively filtered, as discussed below, resulting finally in a set of articles, the vast majority of which satisfies a formal definition of a case series study.

**Figure 1.**
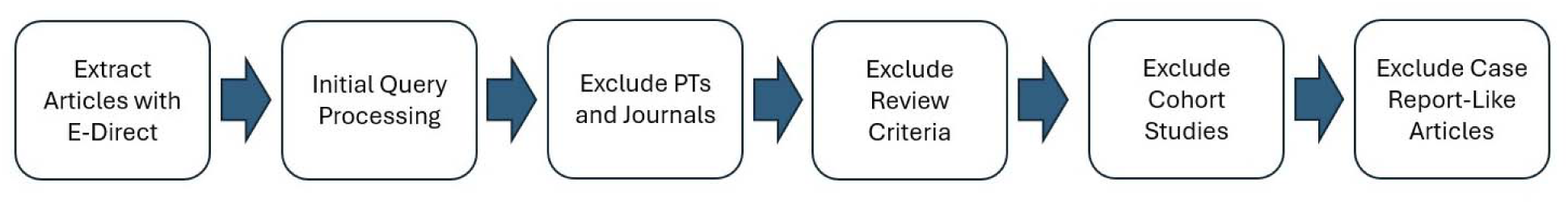
Flowchart of case series articles processing and exclusions. See text for details.

### Preprocessing and feature extraction

The Arrowsmith biomedical tokenizer (https://arrowsmith.psych.uic.edu/arrowsmith_uic/download/tokenizer.txt) was used to lowercase and word tokenize the title and abstract text. To process the article title and abstract text and MeSH terms, the Arrowsmith 1400 word stoplist (https://arrowsmith.psych.uic.edu/arrowsmith_uic/data/stopwords_1400) was implemented, except that the following words were excluded from the stoplist: “case”, “one”, “two”, “three”, “four”, “five.” “Case” was excluded due to the importance of “case series” and “case reports”, and the number words were excluded to allow for later sample size extraction (i.e., number of cases or patients described in the study). The following highly frequent stop words were removed from the MeSH terms: “Humans”, “Female”, “Male”, “Animals”, “Adult”, “Middle Aged”, “Aged”, “Adolescent, Child”, “Rats”, “Mice”, “Time Factors”, “Treatment, Outcome”, “Child, Preschool”, “United States”, “Aged, 80 and over”, “Pregnancy”, “Risk Factors”, “Infant.” Unigrams, bigrams, and trigrams were extracted from the processed article titles and abstracts. N-grams appearing in less than 10 articles or more than 70% of articles were removed. For the remaining n-grams, their document frequency as a percentage of the total articles in each set was calculated. A similarly normalized document frequency was calculated for each MeSH term, publication type, and journal.

Bigrams matching the pattern “X case(s)” or “X patient(s)” where X is a number were extracted and written number words were converted into numerals (e.g., “twenty” into “20”). “COVID-19”, “case series”, “case report”, and “case control” mentions were converted to “[removed COVID-19 mention]”, “[removed case series]”, “[removed case report]”, and “[removed case control]” to avoid erroneously extracting these references as sample sizes. The bigrams were then grouped into ranges of 1-5, 6-10, 11-19, and >= 20 in order to better capture the number of patients or cases discussed by articles. 100 articles were manually evaluated to ensure each article was sorted into the correct category.

### Multi-Tagger model for assigning publication types

Many articles contained in PubMed are not indexed for publication types by NLM at all. In order to estimate the publication types when NLM indexing was not present, we also inferred their assignment to one or more of 50 publication types and study designs as predicted by a probabilistic Support Vector Machine-based machine learning-based model developed by our group, Multi-Tagger [4], which employs metadata features such as title, abstract, journal, MeSH terms, and number of authors. These predictions are publicly viewable for all PubMed articles in the Anne O’Tate value-added PubMed search interface (https://arrowsmith.psych.uic.edu/cgi-bin/arrowsmith_uic/AnneOTate.cgi).

NLM indexing, supplemented by Multi-Tagger predictive scores, were used to identify “case report-like” articles (Figure 2). The case report probability score assigned to each article in the retrieved [title] and [abstract] sets was assigned “case report-like” if either it is indexed as a case report by NLM or predicted by Multi-Tagger using an optimized F1 decision threshold of 0.49 (Figure 2). (The vast majority of NLM-indexed case reports were also predicted to be case reports by Multi-Tagger, confirming that Multi-Tagger scores were accurate for this task.) If neither NLM nor Multi-Tagger predicted the article to be a case report, it was deemed “non-case report-like”.

**Figure 2.**
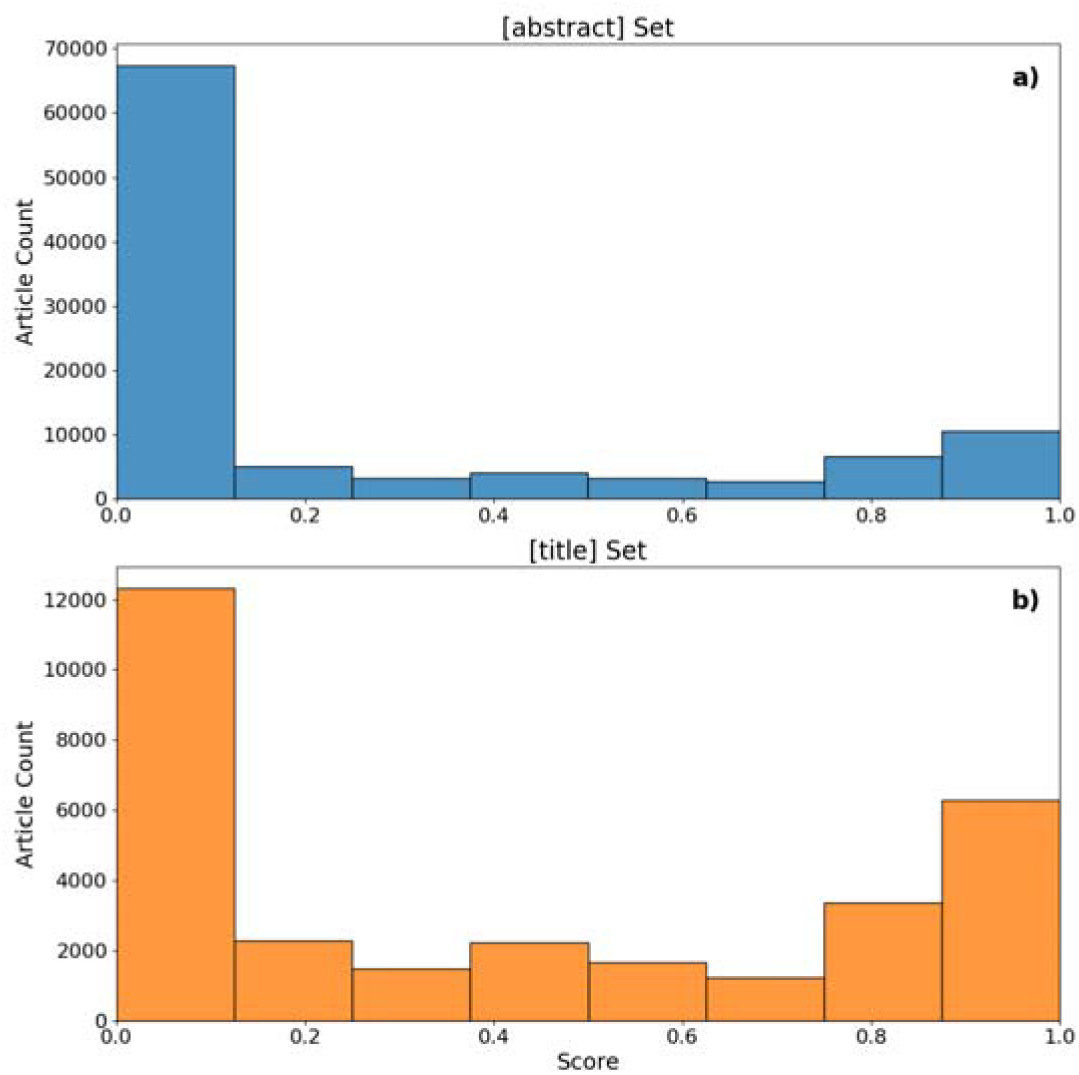
Distribution of Multi-Tagger predictive scores for the case report publication type for both the initial a) [abstract] and b) [title] sets. The optimized F1 score decision threshold was 0.49 assigning articles with a higher score as case report-like and articles with a lower score as non-case report-like.

### Definition of case series article for the purpose of evaluating the proposed training set

Several formal definitions of “case series” were examined, including published definitions [7–9] as well as those proposed by Cochrane [https://community.cochrane.org/sites/default/files/uploads/inline-files/Definitions-Study-Characteritics_Cochrane-COVID-19-Study-Register_0.pdf], Wikipedia [https://en.wikipedia.org/wiki/Case_series], and Sage Research Methods [https://methods.sagepub.com/ency/edvol/encyc-of-epidemiology/chpt/case-reports-case-series]. We settled on a consensus working definition of case series that, while not official, was employed for objectively evaluating and characterizing the articles in the proposed training set:

*A case series is a descriptive study that follows a group of patients who have a similar presenting history, diagnosis, clinical presentation and progression, or prognosis in individual patients, or who are undergoing the same procedure, or share an adverse event, over a certain period of time*.

Several clarifying notes were added for annotators:

- A case series is usually a planned study and generally does not consist of incidental observations.
- A case series is always a group of patients, i.e. at least 3, often 4 or more, but sometimes a large number (e.g., >50).
- A case series has no control group (except for self-controlled studies where the same patient is their own control).
- A case series can satisfy more than one publication type or study design at the same time, e.g. a case series could also incorporate a review, or could also be described as a case report, a cohort study, or even other types of studies within the same article.
- A case series must be observational and cannot be an interventional study. The patients may have undergone a treatment or procedure, and the authors can comment on the efficacy of the treatment or procedure. This may be assessed by e.g., a retrospective chart review. However, if patients are actively recruited for the study, and the aim of the study is to assess clinical outcomes following an intervention, this is an uncontrolled clinical trial instead. The only exception would be if a single article reported a case series study AND a clinical trial study.
- A case series differs from an analytic cohort study insofar as a cohort follows and contrasts TWO different groups of subjects whereas a case series follows one group. However, sometimes a case series compares two subgroups of patients, in which case the distinction between a case series and a cohort study becomes difficult to decide.

## Results

### Retrieving the initial sets of “Case Series” articles and performing initial exclusions

Two article sets, referred to as the [title] set and [abstract] set, were gathered using the PubMed E-Utilities tool to query PubMed and extract articles. The first query gathered articles mentioning the phrase “case series” in the title, while the second focused on articles with “case series” in the abstract only (i.e., not in the title). Both sets included articles published 01/01/1987 - 12/31/2023 and written in English. Each article was downloaded in XML format and the PMID, article title, author last names, abstract text, publication year, publication types, MeSH terms, journal title, journal ISO abbreviation, and page numbers were extracted. Articles indexed according to the following NLM Publication Types and MeSH study designs either in PubMed or Multi-Tagger were immediately excluded for both [title] and [abstract] sets as being inherently inconsistent with being a case series study: “Published Erratum”, “Retraction of Publication”, “Retracted Publication”, “Duplicate Publication”, “Bibliography”, “Portrait”, “Legal Case”, “Lecture”, “Congress”, “Pictorial Work”, “Newspaper Article”, “Book Illustrations”, “Webcast”, “Video Audio Media”, “Electronic Supplementary Materials”, “Comment”, “Editorial”, “Case Control Studies”, “Clinical Trial”, “Controlled Clinical Trial”, “Randomized Controlled Trial”, “Clinical Trial, Phase I”, “Clinical Trial, Phase II”, “Clinical Trial, Phase III”, “Clinical Trial, Phase IV”, “Clinical Trial Protocol”, “Pragmatic Clinical Trial”, “Clinical Trial, Veterinary”, and “Randomized Controlled Trial, Veterinary.”

We also excluded articles published in the following methodological or statistical journals, as likely to be discussing methodology rather than presenting results of a case series study: “Ann Epidemiol”, “Bioinformatics”, “Biom J”, “Biostatistics”, “BMC Genet”, “BMC Med Genomics”, “BMC Med Res Methodol”, “BMC Syst Biol”, “Br J Math Stat Psychol”, “Bull Math Biol”, “Clin Trials”, “Comput Methods Programs Biomed”, “Contemp Clin Trials”, “Control Clin Trials”, “Epidemiology”, “Eur J Epidemiol”, “Genet Sel Evol”, “IEEE ACM Trans Comput Biol Bioinform”, “IEEE Trans Pattern Anal Mach Intell”, “IMA J Math Appl Med Biol”, “Int J Health Geogr”, “Int J Methods Psychiatr Res”, “J Anim Breed Genet”, “J Biopharm Stat”, “J Clin Epidemiol”, “J Pharmacokinet Pharmacodyn”, “J R Soc Interface”, “Lifetime Data Anal”, “Math Biosci”, “Math Med Biol”, “Mol Syst Biol”, “Neural Netw”, “Pharm Stat”, “Phys Rev E Stat Nonlin Soft Matter Phys”, “PLoS Comput Biol”, “Psychol Methods”, “Stat Appl Genet Mol Biol”, “Stat Med”, “Stat Methods Med Res”, and “Theor Biol Med Model”.

Since case series studies often include a review of the literature, articles indexed as reviews were not removed initially. However, at this point, articles mentioning both terms “systematic review” and “reports” in the title were removed from the [title] set, as these were comprised of systematic reviews covering case reports and case series.

### “Case series” [title] set

The initial set of 33,720 articles mentioning “case series” in the title was trimmed by removing those without abstracts, with incompatible publication types, those published in methodological and statistical journals, and those with “reports” and “systematic reviews” in the title, leaving 28,829 articles in the initial set to be characterized further.

A striking finding is that more than half of these articles are indexed as case reports by NLM or predicted by the Multi-Tagger model. In fact, in the [title] set, the distribution of Multi-Tagger predictive scores for the Case Reports publication type was clearly bimodal (Figure 2). Of note, 8 of the top 20 journals publishing case report-like articles explicitly had “case reports” in the title of the journal (vs. only 1 of the top 20 in the other subset).

Case report-like (i.e., NLM-indexed or predicted to be Case Reports) and non-case-report like (i.e., all others) subsets were characterized further in terms of the number of patients studied. Statements of the form “X cases” and “X patients” mentioned in the title or abstract were extracted separately, where X is a number written either as a numeral or as a word. As shown in Table 1, in the [title] set, 44.7% of the case report-like articles mentioned 5 or fewer cases or patients in the abstract, vs. 12.9% of the non-case report like subset. Conversely, only 13.2% of the case report-like articles mentioned 11 or more cases or patients in the abstract, vs. 42.3% of the non-case report-like subset (Table 1).

**Table 1.** Abstract mentions of “X cases” and “X patients” in the [title] and [abstract] sets, separated into case report-like and non-case report-like subsets.

|  | Case Report-Like |  | Non-Case Report-Like |  |
| --- | --- | --- | --- | --- |
|  | [title] Set | [abstract] Set | [title] Set | [abstract] Set |
| 1-5 case(s) | 3,162 (20.57%) | 1,243 (11.93%) | 436 (3.45%) | 659 (1.72%) |
| 1-5 patient(s) | 3,705 (24.10%) | 2,841 (27.27%) | 1,189 (9.42%) | 2,579 (6.71%) |
| 6-10 case(s) | 670 (4.36%) | 268 (2.57%) | 335 (2.65%) | 440 (1.15%) |
| 6-10 patient(s) | 1,521 (9.89%) | 1,119 (10.74%) | 1,565 (12.40%) | 2,975 (7.74%) |
| 11-19 case(s) | 253 (1.65%) | 100 (0.96%) | 272 (2.16%) | 422 (1.10%) |
| 11-19 patient(s) | 599 (3.90%) | 433 (4.16%) | 1,470 (11.65%) | 3,683 (9.59%) |
| >=20 case(s) | 402 (2.61%) | 161 (1.55%) | 594 (4.71%) | 1,543 (4.02%) |
| >=20 patient(s) | 779 (5.07%) | 568 (5.45%) | 3005 (23.81%) | 13,138 (34.20%) |
| None | 4,283 (27.86%) | 3,686 (35.38%) | 3,755 (29.75%) | 12,976 (33.78%) |

Several previous analyses have pointed out that many case series articles satisfy study design criteria of descriptive and/or population-based cohort studies [7–9]. However, we found that only 5.0% of the non-case report-like articles were indexed by NLM or predicted by Multi-Tagger as Cohort Studies, and only 2.51% were indexed as Clinical Study. Other clinical publication types were equally or more uncommon (<1%), e.g., Evaluation Study, Predictive Value of Tests, Cross-Sectional Studies, and Longitudinal Studies. This argues that the non-case report-like articles comprise a unique type of biomedical literature, not well covered by any existing indexing terms.

### “Case series” [abstract] set

The set of articles mentioning “case series” in the title or abstract (106,033 articles) was trimmed by removing articles with “case series” in the title, incompatible publication types, and those published in methodological and statistical journals, leaving 66,861 articles in the set to be characterized further. About 10% of the articles were case report-like (Figure 2). Similar to the [title] set, almost half of the non-case report-like articles in the [abstract] set mentioned >10 cases or patients (Table 1).

### Manual evaluation of the provisional corpus

At this point, we contemplated combining the non-case report-like [title] and [abstract] sets as a representative corpus for indexing case series studies. However, it was unclear whether the great majority of these articles would satisfy formal definitions of “case series”. Although there is no official, entirely accepted or consistent definition of “case series” [8], we examined several of the available definitions and created our own consensus definition to guide manual annotation (see Methods). We randomly chose 50 articles from the non-case report-like “case series” [title] subset and 50 from the “case series” [abstract] articles. We also added 10 randomly chosen case report-like articles, to learn whether these would also satisfy definitions of case series. These 110 articles were shuffled and presented to two annotators, blind to article assignment, who independently read the title and abstract (and full-text if necessary), scoring whether it satisfied our formal working definition of “case series”, noting the type of design and any unusual features, and extracting the total number of patients studied. Differences were reconciled by discussion.

Of the 50 “case series” [title] articles, 88% were judged to satisfy the working definition. Three prominent subtypes of case series were detected: One subtype followed a group of patients who shared a given diagnosis or condition; one followed a group of patients who were subjected to a particular type of surgery or other treatment or intervention; and a third carried out anatomical or technical measurements on samples derived from subjects. Most studies were retrospective, ranging from three to thousands of patients. A few studies, which had both prospective design and active recruitment of patients, were regarded as uncontrolled clinical trials and scored as not a case series study. A few articles compared subjects to control groups, which also excluded them from our definition. Note that of the 10 case report-like articles evaluated, 9 did satisfy the definition of “case series” as well. Of the two articles that were predicted to be cohort studies according to the Multi-Tagger model, both satisfied the criteria to be called case series as well. Thus, the scope of case series studies vs. case reports is not entirely distinct, and a single article may show more than one type of design.

Among the 50 “case series” [abstract] articles, the same range of study heterogeneity was observed as in the [title] set. However, 15 of the articles were systematic reviews or other reviews (not reporting the results of an individual case series study) that had not been adequately removed by previous rules. Apart from these reviews, 94.2% of the [abstract] articles were judged to satisfy the working definition of case series. We then tested an additional exclusion rule, to remove any articles from the [abstract] set that were indexed as Review or Systematic Review by NLM, or predicted as such by Multi-Tagger. Since this rule removed 14 of the 15 reviews from the test sample but none of the other articles, the rule was thus implemented across the entire [abstract] set.

After the evaluation of the 110 articles, we implemented the review exclusion rule in the [abstract] set (as just discussed), further removed any articles indexed as Cohort Studies by NLM or predicted by Multi-Tagger in the combined set, and finally removed the case-report like articles. The final proposed corpus thus consisted of 12,621 [title] articles and 38,415 [abstract] articles, or a total combined of 51,036 case series studies. The corpus has been deposited in the UIC INDIGO data repository (https://indigo.uic.edu/, https://doi.org/10.25417/uic.28593611.v1).

### Automated assignment of Case Series to PubMed articles

Recently, we have developed a series of transformer models that assigns predictive scores for up to 72 publication types and study designs (collectively referred to as PTs), which improves upon the earlier SVM-based Multi-Tagger model. The queries used to generate most of the training labels, the model architecture, and the input features are described [10]. Briefly, this model uses WeighCon supervised contrastive learning, and title, abstract, and various metadata-based features, such as journal name, served as input into the model. Training examples were taken from PubMed records in English published between 1987 and 2023 that had been manually assigned PTs by NLM, adjusting the number of examples per PT by under-sampling to deal with the fact that certain PTs have many more articles than others [5]. Using this same transformer model architecture, we added new PTs including “Case Series”, by incorporating the cleaned-up corpus described above with the data for other PTs. Overall, this model was developed using a stratified random sample of 1,284,378 articles, including 19,721 case series studies included in our corpus, which was stratified by PT to preserve label distribution. The dataset was split using 70/10/20 train/validation/test splits, again stratified to preserve label distribution in each split. Articles were tagged if the probabilistic prediction of the model was above a threshold, which was determined empirically to optimize for F1 within the validation set. We further added a post-model exclusion rule to remove articles that mentioned “case series” in the abstract only, that were predicted to be Review or Systematic Review.

The test set consisted of 256,897 articles, among which were 3,944 case series articles included in our corpus but not used for the training or validation of the model. Case series performance of this model was excellent (precision = 0.887, recall = 0.952, F1 = 0.918, PR-AUC = 0.941). To test generalizability and performance under real-world conditions, the model was applied to all PubMed articles published in 2024, and a binary assignment of Yes or No was given to predicted case series articles using the threshold optimized on the validation set. Of the 1,723,429 articles in the 2024 PubMed set that were processed by the model, only 9,450 (0.5%) were tagged as Case Series, the overwhelming majority of which mentioned the phrase “case series” in title or abstract. Conversely, 765 articles mentioned that phrase but were NOT predicted to be Case Series, indicating that the model did not simply use the presence of that phrase as a deciding feature. A few (n = 33) articles did not mention the exact phrase “case series” but were tagged as Case Series; about half of these mentioned some lexical variant such as “cases series”, while others used alternative informative phrases such as “retrospective chart review”.

A random sample of 100 articles tagged as Case Series by the model were manually examined by two investigators under blind conditions, reconciling any disagreements. Of the 100 articles, when the title and abstract of the article was screened, 95% satisfied our formal definition of case series. The remaining 5% comprised clinical trials, a case-control study, and a review of case series studies. Only 4 of the 100 articles were indexed as Case Report by NLM. However, when the full-text of the article was examined, we discovered that 7 of the case series that involved prospective design and interventions were judged to be clinical trials instead, giving overall accuracy of 88%. None of the 100 articles mentioned trial registry numbers (e.g., NCT numbers from ClinicalTrials.gov).

## Discussion

### Corpus of articles mentioning “case series” in title or abstract

As shown in Table 2, only about half of the articles that mention “case series” in the title or abstract are actually typical case series studies. Almost half of articles that mention the phrase “case series” in title strongly resemble case reports, both because they are explicitly indexed as such by NLM (and/or predicted as such by our publication type model Multi-Tagger [4]), and because they predominantly deal with a small number of patients. Although they generally satisfied our formal working definition of case series, indicating that case series and case reports are not entirely exclusive concepts [11], we removed case report-like articles from our final case series corpus, with the rationale that they are better indexed as the existing standard Case Reports Publication Type.

**Table 2.** Estimates of article types as a percentage of all retrieved articles mentioning “case series” in the title or in the abstract, based on the exclusion procedures described in this paper.

|  | [title] set | [abstract] set | Sum | percentage |
| --- | --- | --- | --- | --- |
| Methodology | 46 | 128 | 174 | 0.2% |
| Review | 90 | 15593 | 15683 | 15.1% |
| Case Report | 15374 | 10419 | 25793 | 24.9% |
| Case Series | 12621 | 38415 | 51036 | 49.2% |
| Other | 5589 | 5381 | 10970 | 10.6% |
| Sum | 33720 | 69936 | 103656 | 100% |

Another surprising finding is that very few articles that mention the phrase “case series” are indexed as Cohort Studies by NLM, nor predicted as such by Multi-Tagger. Several prior analyses have pointed out that many, perhaps most case series also share aspects of design with descriptive and/or population-based cohort studies [7–9], wherein a single group of subjects are followed over time, either after a particular diagnosis was made or after a particular intervention was carried out. However, even if case series do share general features with descriptive and population-based cohort studies, these stand in contrast to classical analytic cohort studies which compare two groups or subsets of patients [12].

### Automated assignment of Case Series to PubMed articles

The cleaned-up corpus of case series articles was employed for training, validation, and testing of a transformer based model that assigned predictive scores for 72 different publication types and study designs, including a new label of “case series”. Performance of the model was excellent, with formal evaluation indicating F1 = 0.918 and manual assessment showing 88% accuracy.

### Limitations

Our scheme has three obvious limitations: First, we do not know the true prevalence of case series articles within the overall biomedical literature. We suspect that most authors do employ the phrase “case series” in writing up case series studies, and the transformer model was able to detect some articles that employed alternative phrases such as “retrospective chart review”, however, we acknowledge that that the current corpus may be missing some relevant articles. A second limitation is that the transformer model did not examine features within the full text of articles, especially the Methods section. Indeed, we found that a small percentage of prospective case series that were tagged by the model were actually revealed to be clinical trials once the full-text was examined. We are currently evaluating transformer models that do incorporate full text features. Third, there is no official or universally agreed-upon definition of a case series study. We have created a consensus definition that should suffice for most users and use cases, but we acknowledge that there is not always a hard distinction between a case series and other designs (case reports, cohort studies and uncontrolled clinical trials). The probabilistic output of the transformer model may partially help in this regard, as it should give high-confidence predictive scores to “typical” case series articles and lower scores to those which are atypical.

## Conclusion

Our analysis indicates that no existing publication type or study design indexing term captures typical case series studies well, supporting our effort to create a corpus and create a new specific “case series” indexing term. In the near future, once the transformer model has been optimized across all publication types and study designs, the model scores for Case Series will be processed across all PubMed articles and disseminated publicly for the use of the biomedical community.

As well, we will create a free, public API so that users can input article records or full-text articles (including non-PubMed articles) and receive predictive scores for the transformer model in real time.

## Data Availability

The case series training corpus has been deposited in the UIC INDIGO data repository (https://indigo.uic.edu/ https://doi.org/10.25417/uic.28593611.v1). The transformer model to assign predictive scores is previously described [10] and is archived at https://github.com/ScienceNLP-Lab/MultiTagger-v2.

## Author Contributions

AS: Data curation, formal analysis, Writing – original draft.

AWH: Formal analysis, Investigation, Writing – review & editing.

AMT: Formal analysis, Investigation.

JDM: Methodology, Software, Validation.

NRS: Conceptualization, Funding acquisition, Methodology, Supervision, Writing – review & editing.

## References

1. Murad MH, Sultan S, Haffar S, Bazerbachi F. Methodological quality and synthesis of case series and case reports. BMJ Evid Based Med. 2018 Apr;23(2):60–63. doi: 10.1136/bmjebm-2017-110853.

2. Smith EG, Patel KM. The Role of Case Series and Case Reports in Evidence-Based Medicine. J Clin Psychopharmacol. 2024 Mar-Apr 01;44(2):81–85. doi: 10.1097/JCP.0000000000001826.

3. Preskorn SH, Armstrong AG. Can the Publication of Case Series or Case Reports Lead to a Change in Clinical Practice? J Psychiatr Pract. 2023 Mar 1;29(2):137–141. doi: 10.1097/PRA.0000000000000701.

4. Cohen AM, Schneider J, Fu Y, McDonagh MS, Das P, Holt AW, Smalheiser NR. Fifty ways to tag your pubtypes: Multi-tagger, a set of probabilistic publication type and study design taggers to support biomedical indexing and evidence-based medicine. medRxiv. 2021 Jul 16:2021–07. doi: 10.1101/2021.07.13.21260468.

5. Menke JD, Kilicoglu H, Smalheiser NR. Publication Type Tagging using Transformer Models and Multi-Label Classification. AMIA Annu Symp Proc. 2025 May 22;2024:818–827.

6. Wallace BC, Noel-Storr A, Marshall IJ, Cohen AM, Smalheiser NR, Thomas J. Identifying reports of randomized controlled trials (RCTs) via a hybrid machine learning and crowdsourcing approach. J Am Med Inform Assoc. 2017 Nov 1;24(6):1165–1168. doi: 10.1093/jamia/ocx053.

7. Sargeant JM, O’Connor AM, Cullen JN, Makielski KM, Jones-Bitton A. What’s in a Name? The Incorrect Use of Case Series as a Study Design Label in Studies Involving Dogs and Cats. J Vet Intern Med. 2017 Jul;31(4):1035–1042. doi: 10.1111/jvim.14741.

8. Esene IN, Ngu J, El Zoghby M, et al. Case series and descriptive cohort studies in neurosurgery: The confusion and solution. Childs Nerv Syst 2014;30:1321–1332.

9. Dekkers OM, Egger M, Altman DG, et al. Distinguishing case series from cohort studies. Ann Intern Med 2012;156:37–40.

10. Menke JD, Ming S, Radhakrishna S, Kilicoglu H, Smalheiser NR. Enhancing automated indexing of publication types and study designs in biomedical literature using full-text features. medRxiv. 2025 Apr 25:2025–04. 10.1101/2025.04.23.25326300

11. Abu-Zidan FM, Abbas AK, Hefny AF. Clinical “case series”: a concept analysis. Afr Health Sci. 2012 Dec;12(4):557–62.

12. “Cohort Studies”[MeSH Terms] https://www.ncbi.nlm.nih.gov/mesh/?term=cohort+studies.

